# Association of Daily Step Count and Postoperative Surgical Complications Among All of Us Research Participants

**DOI:** 10.1101/2023.12.08.23299235

**Authors:** Carson J. Gehl, Nathaniel B. Verhagen, Tahseen J. Shaik, Kaitlyn Nimmer, Xin Yang, Yun Xing, Bradley W. Taylor, Mochamad M. Nataliansyah, Sarah L. Kerns, Anai N. Kothari

**Affiliations:** Department of Surgery, Division of Surgical Oncology, Medical College of Wisconsin, Milwaukee, WI; Department of Biostatistics, Medical College of Wisconsin, Milwaukee, WI; Clinical and Translational Science Institute of Southeastern Wisconsin, Medical College of Wisconsin, Milwaukee, WI; Department of Radiation Oncology, Medical College of Wisconsin, Milwaukee, WI; Data Science Institute, Medical College of Wisconsin, Milwaukee, WI

**Author notes:** Corresponding Author: Anai N. Kothari, MD MS FSSO, Medical College of Wisconsin, Division of Surgical Oncology, 8701 W Watertown Plank Road, Milwaukee, Wisconsin 53226, USA, (414)955-1443. **Meeting Presentation:** American College of Surgeons (ACS) Clinical Congress, Boston, MA, October 2023.

## Abstract

**Background:** The association between preoperative wearable device step counts and surgical outcomes has not been examined using commercial devices linked to electronic health records (EHR). This study measured the association between daily preoperative step counts and postoperative complications.

**Study Design:** Data was obtained using the All of Us (AOU) Research program, a nationwide initiative to collect EHR and health-related data from the population. Included were patients who underwent a surgical procedure included in the National Surgical Quality Improvement Program (NSQIP) targeted procedures dataset. Excluded were patients who did not have available physical activity FitBit data. Primary outcome was the development of a postoperative complication. All analyses were performed in the AOU researcher workbench.

**Results:** Of 27,150 patients who underwent a surgical procedure, 475 participants with preoperative wearable data were included. 74.7% were female and 85.2% were White. The average age was 57.2 years. The overall rate of postoperative complications was 12.6%. Patients averaging fewer than 7,500 daily steps were at increased odds for developing a postoperative complication (OR 1.83, 95% CI [1.01, 3.31]). Following adjustment for age, sex, race, comorbid disease, body mass index (BMI), and relative procedure risk, patients with a baseline average steps/day < 7,500 were at increased odds for postoperative complication (aOR = 2.06, 95% CI [1.05, 4.06]).

**Conclusions:** This study found an increase in overall postoperative complication rate in patients recording lower average preoperative step counts. Patients with a baseline of less than 7,500 steps per day had increased odds of postoperative complications in this cohort. This data supports the use of wearable devices for surgical risk stratification and suggests step count may measure preoperative fitness.

## INTRODUCTION

Risk stratification is a critical part of successful perioperative care and guides efforts for optimization when preparing a patient for surgery.^1, 2^ In recent years, preoperative fitness assessment has gained increasing recognition as a tool for surgical risk stratification. Cardio-pulmonary exercise testing (CPET) can be used to test patient fitness and plan postoperative care based on individual risk.^3^ Some have also worked to implement subjective, self-reported measures to more easily estimate functional capacity prior to surgery.^4^

Beyond planning postoperative care, recent data demonstrates a benefit for prehabilitation, which focuses on improving patient fitness prior to surgery by targeting modifiable lifestyle factors such as diet, exercise, smoking, and mental health.^5^ Interventional studies targeting physical activity in the preoperative period have demonstrated the ability to improve functional capacity and postoperative outcomes.^6–8^ Tracking patient fitness and physical activity in the preoperative period may facilitate the identification of modifiable targets to improve surgical outcomes.

Despite these advances, challenges remain in the clinical implementation of these approaches. Traditional methods of functional capacity assessment have depended on CPET or subjective/self-reported measures. While CPET offers a robust measure of fitness and can inform perioperative care, it requires an investment of substantial resources.^9^ It is unclear how accurately subjective assessment portrays patient fitness.^10, 11^ Wearable devices, such as Fitbits, provide a wealth of objective physical activity data and could offer a convenient, cost-effective, and accessible way to inform clinicians about patients’ overall fitness over an extended period. Recently, several studies have examined the association of preoperative step counts recorded by wearable pedometer devices with postoperative outcomes.^12–15^ However, prior work has typically been confined to small prospective cohorts undergoing a single type of procedure. In addition, no study utilizing a large-scale EHR dataset has measured the association between step counts and postoperative outcomes across procedures of varying organ sites and complexity.

The All of Us (AOU) Research Program is a National Institutes of Health (NIH)-funded effort to collect EHR, health questionnaire, genetic, and digital health technology data from over 1 million people in the United States.^16^ Participants have the opportunity to share wearable device data, providing physical activity data that can be linked to their EHR data. The primary aim of this study was to investigate the association between preoperative daily step counts and postoperative adverse outcomes in AOU participants who have undergone elective surgical procedures.

## METHODS

### Study Participants

This was a retrospective cohort study using the All of Us Research Program Controlled Tier version 7 data set and included participants who contributed wearable device physical activity data along with EHR data. Participants 18 and older can be enrolled in the AOU Research Program after an informed consent process at an AOU-associated clinic. The AOU Research Program was approved by the AOU Institutional Review Board, and additional IRB approval is not required for studies using AOU data through the Researcher Workbench, a secure cloud-based platform. Deidentified data was accessed and analyzed within the AOU Researcher Workbench by authors who had completed AOU Responsible Conduct of Research training. Included were patients who underwent an elective surgical procedure defined using linked participant EHR data and source provided Current Procedural Terminology (CPT) codes. The selection of CPT codes for study inclusion was based on the same criteria used by the National Surgery Quality Improvement (NSQIP) program’s Procedure Targeted dataset (Supplementary Table 1). NSQIP data is not available in AOU. For patients who underwent multiple procedures, only the most recent was included for analysis. Participants with no available physical activity data prior to surgery were excluded.

### Independent Variables

Age, sex, and race were obtained from AOU participant surveys. The most recent body mass index (BMI) measurement before the procedure was obtained from participant EHR. For participants without a BMI measurement (missingness 37.6%), the median was imputed. A sensitivity analysis was performed without BMI, BMI included without imputation, and median imputation, and no difference was noted based on approach. The BMI data showed a non-normal distribution, making the median a more robust measure than the mean. Additionally, the missing data were missing at random, supporting the use of a simple imputation method. Procedure risk was determined using the median length of stay (LOS) by CPT code. Procedures with median LOS of 0-1 days were deemed low-risk, 2-4 days medium-risk, and 5+ days high-risk. Comorbid disease was defined using ICD codes in participant EHR prior to surgery, indicating any condition defined under the Elixhauser comorbidity index. Physical activity was measured by calculating the mean daily steps for participants who had any preoperative physical activity data. Fitbit devices are not provided to AOU participants. Instead, participants who already own Fitbits can share their data with AOU. Fitbits have been utilized in many studies aimed at tracking patient activity, and prior studies have shown they provide reliable step counts.^17, 18^ While there is likely some inherent error in device recording, these devices have been proven to be effective and reliable for research purposes, particularly in their measurement of steps. The mean steps/day was chosen for analysis as this is what is typically provided by digital health applications to consumers, thus making results more applicable to users.

### Development of Postoperative Complications

The primary outcome of the study was the development of any adverse event within 90 days of surgery. Adverse events included pneumonia, respiratory failure, pulmonary embolism, sepsis, cardiac arrhythmia, renal failure, urinary tract infection, and deep vein thrombosis. Systemized Nomenclature of Medicine (SNOMED) concepts were used for defining postoperative adverse events (Supplementary Table 2). This composite outcome was selected based on prior studies using similar large, EHR-based datasets to investigate surgical outcomes.^19^ Mortality was not included as a primary outcome as there were no deaths that occurred during the postoperative period in this cohort.

### Statistical Analysis

Descriptive statistics summarizing participant demographic and other clinically relevant variables were represented by median and IQR for continuous variables and frequency for categorical variables. Two-sample t-tests and chi-squared tests were performed for normally distributed continuous and categorical variables, respectively, to compare demographic and clinically relevant data between participants in stratified groups. Adjusted analyses were conducted with multivariable logistic regression. All analyses were conducted in the AOU Researcher Workbench using Jupyter Notebook software version 6.4.8. The following packages were used for analysis: pandas, os, statsmodels, numpy, matplotlib, scipy, seaborn, and datetime. Participant counts of less than 20 are not reported to comply with the AOU Data and Statistics Dissemination Policy.

## RESULTS

### Descriptive Statistics

475 patients met our inclusion criteria. Demographic and clinical information about the cohort is provided in Table 1. Of the included participants, 60 (12.6%) experienced an adverse event within 90 days following their procedure. Patients who experienced an adverse event were more likely to be male (43.3% vs. 19.8%; P < 0.001), have 4+ comorbid conditions (76.7% vs. 11.6%; P < 0.001), and underwent a medium-or high-risk procedure (41.7% vs. 21.9%; P < 0.001).

**Table 1:** Baseline characteristics of study cohort.

| **Patient Characteristics (N = 475)** |  |
| --- | --- |
| Preoperative steps/day |  |
| **Median, (Q1, Q3)** | 6673 (4770, 9140) |
| Days of preoperative activity recorded |  |
| **Median, (Q1, Q3)** | 700 (240, 1343) |
| Age, years |  |
| **Median, (Q1-Q3)** | 59.3 (48.0, 67.4) |
| **18-29** | < 20 (< 4.2%) |
| **30-49** | > 112 (> 23.6%) |
| **50-64** | 174 (36.6%) |
| **65+** | 169 (35.6%) |
| Sex at birth |  |
| **Female** | 355 (74.7%) |
| **Male** | > 100 (> 21.1%) |
| **Missing or Unknown** | < 20 (< 4.2%) |
| Race |  |
| **White** | 405 (85.3%) |
| **Black or African American** | 23 (4.8%) |
| **Other or Unknown** | 47 (9.9%) |
| Ethnicity |  |
| **Not Hispanic or Latino** | 433 (91.2%) |
| **Hispanic or Latino** | 22 (4.6%) |
| **Other or Unknown** | 20 (4.2%) |
| Elixhauser Comorbidity Index |  |
| **0** | 67 (14.1%) |
| **1** | 56 (11.8%) |
| **2** | 80 (16.8%) |
| **3** | 64 (13.5%) |
| **4+** | 208 (43.8%) |
| Body Mass Index (BMI) |  |
| **Median (Q1, Q3)** | 29.7 (25.2, 35.9) |
| Comorbidities |  |
| **Hypertension, Uncomplicated** | 210 (44.2%) |
| **Obesity** | 166 (34.9%) |
| **Solid Tumor Without Metastasis** | 142 (29.9%) |
| **Depression** | 139 (29.3%) |
| **Chronic Pulmonary Disease** | 125 (26.3%) |
| **Hypothyroidism** | 96 (20.2%) |
| **Diabetes, Uncomplicated** | 67 (14.1%) |
| **Liver Disease** | 59 (12.4%) |
| Relative Risk of Surgery |  |
| **Low Risk** | 359 (78.6%) |
| **Medium Risk** | 90 (19.7%) |
| **High Risk** | 26 (5.7%) |

### Exploratory analysis to determine step threshold

Several step thresholds (average steps/day) were tested to determine observed differences in the rate of postoperative events. At increments of 500 steps, patients were stratified into two groups: active and inactive. For each threshold, the adverse event rate was calculated for those above (i.e. active) and below (i.e. inactive) the average daily step count (Figure 1). Further, univariable logistic regression models were performed to examine the relationship between step counts and odds of postoperative complication (Figure 2). Based on this, a preoperative activity level of 7,500 steps/day was used for further analyses. This point was chosen as it displayed a strong association with the primary outcome and because prior studies have shown step counts of 7000-8000 steps/day to be indicative of moderate to vigorous physical activity and are associated with reduced mortality. Additionally, dividing the cohort at 7500 steps achieved a balanced sample size between the two groups, and thus represents a realistic and pragmatic activity goal that could set for patients in a real clinical setting.^20, 21^ Baseline characteristics of participants in the active vs. inactive group are shown in Supplementary Table 3.

**Figure 1:**
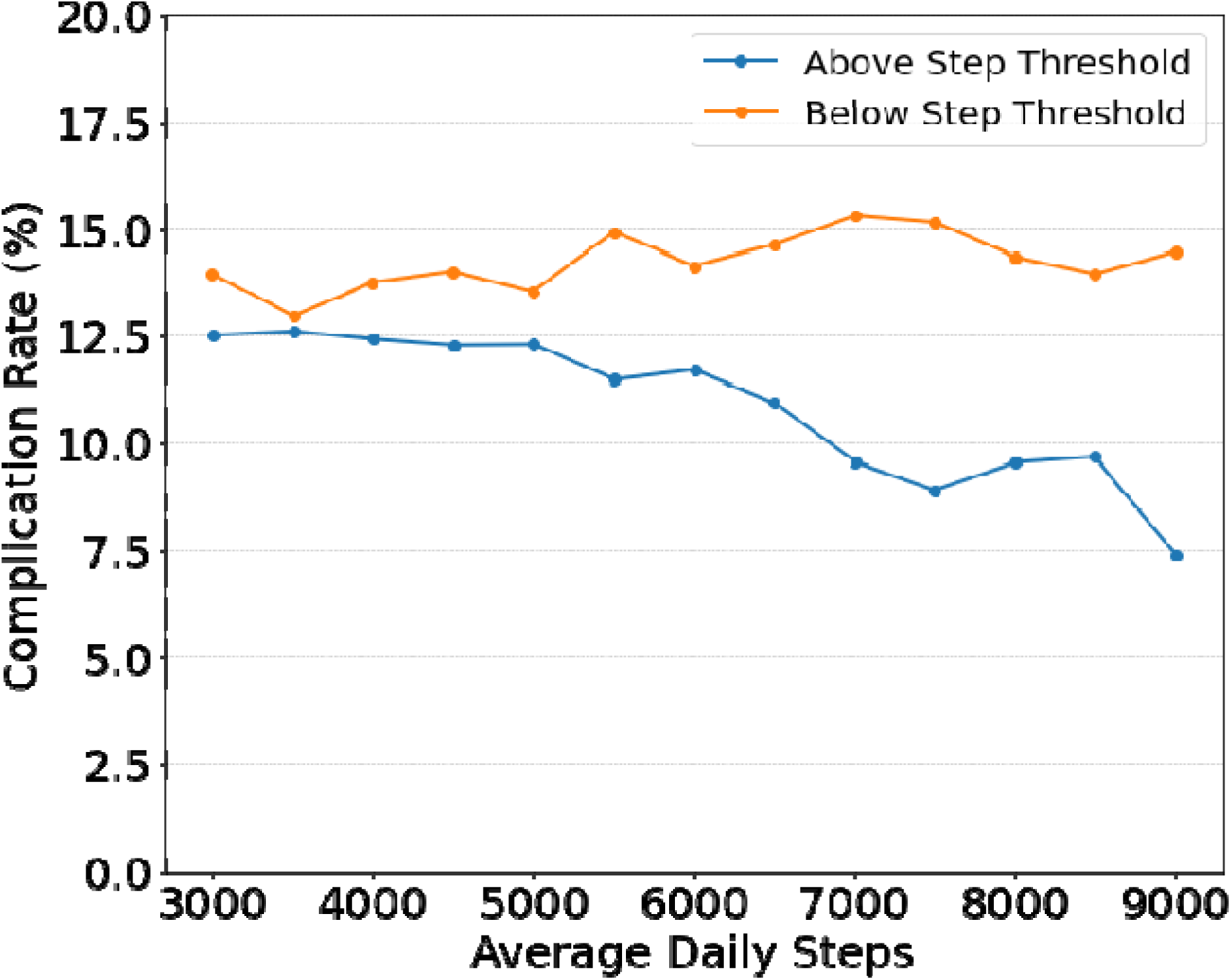
Average complication rate (any adverse event) above and below daily step count threshold, in 500 step increments.

**Figure 2:**
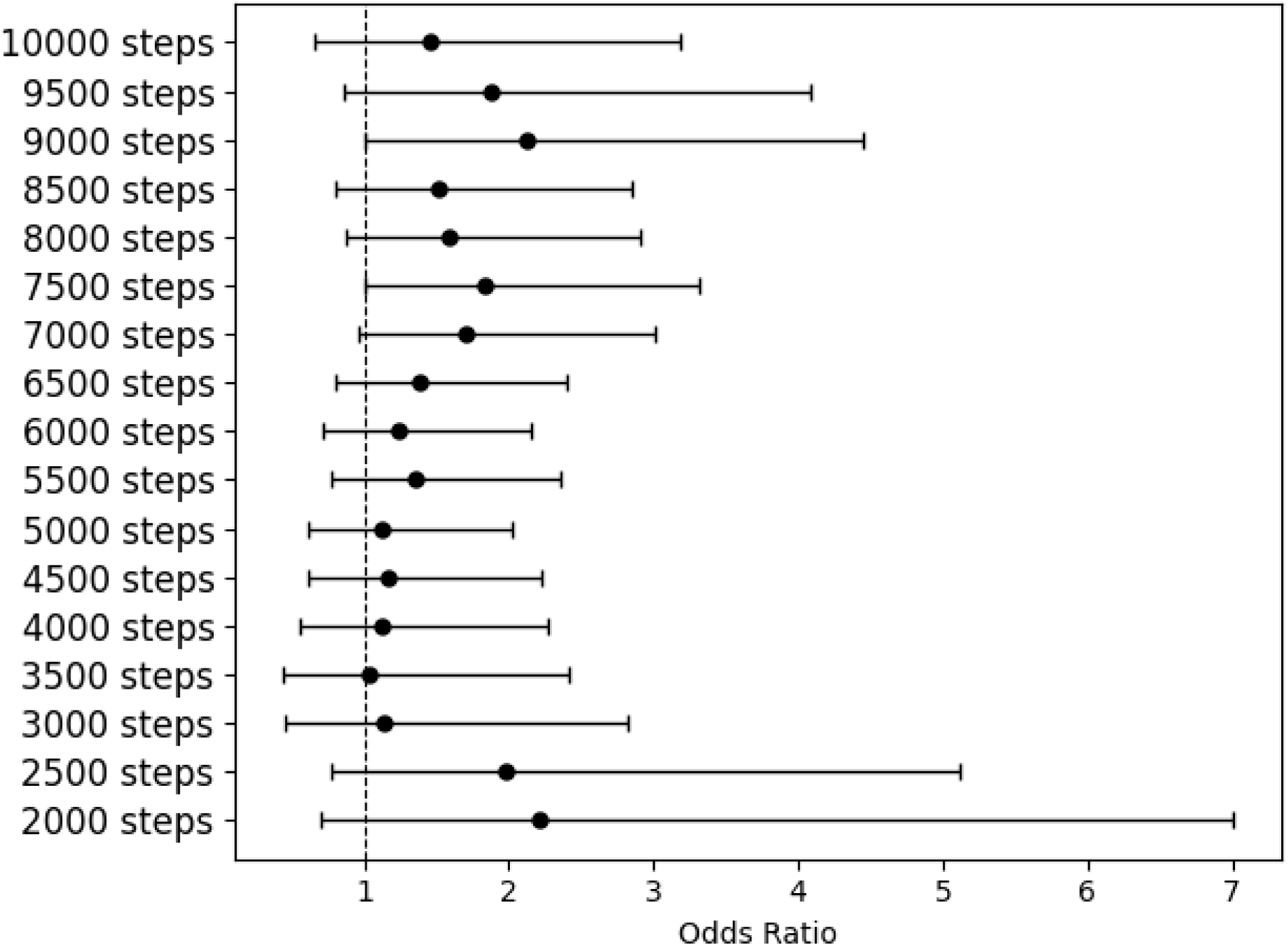
Association between average daily steps (< y-axis) and development of a postoperative complication based on unadjusted logistic regression model. Based on these findings, a threshold of 7500 steps was utilized for further analyses.

### Preoperative step count and 90-day postoperative adverse outcomes

On unadjusted analysis, the rate of 90-day postoperative adverse events was greater in patients who had recorded fewer than 7,500 daily steps prior to their procedure than those who averaged 7,500 or more steps a day (OR 1.83 [1.01, 3.31]). Given the relatively low event rate, there was insufficient evidence to conclude that individual complications were associated with the 7,500-step threshold. For example, amongst those in the inactive group, the odds of deep vein thrombosis were 2.03 [0.21, 19.65], surgical site infection 1.69 [0.33, 8.82], respiratory failure 2.03 [0.21, 19.65], urinary tract infection 1.88 [0.59, 6.01], cardiac arrhythmia 1.45 [0.66, 3.14], renal failure 1.12 [0.27, 4.76], sepsis 0.67 [0.09, 4.80], pulmonary embolism 2.03 [0.21, 19.65], and pneumonia 0.22 [0.02, 2.14] compared to the active group (Supplementary Figure 1).

Following adjustment for age, sex, race, comorbid disease, body mass index (BMI), and relative procedure risk, patients with less than 7,500 average daily preoperative steps had increased odds of 90-day postoperative complications (aOR 2.06 [1.05, 4.06]; Figure 3). A final fitted model was used to calculate individual-level probability of developing a postoperative complication. A higher average probability of postoperative complications was observed for those taking <7,500 steps/day (15.1% vs. 8.9%; P < 0.001; Figure 4).

**Figure 3:**
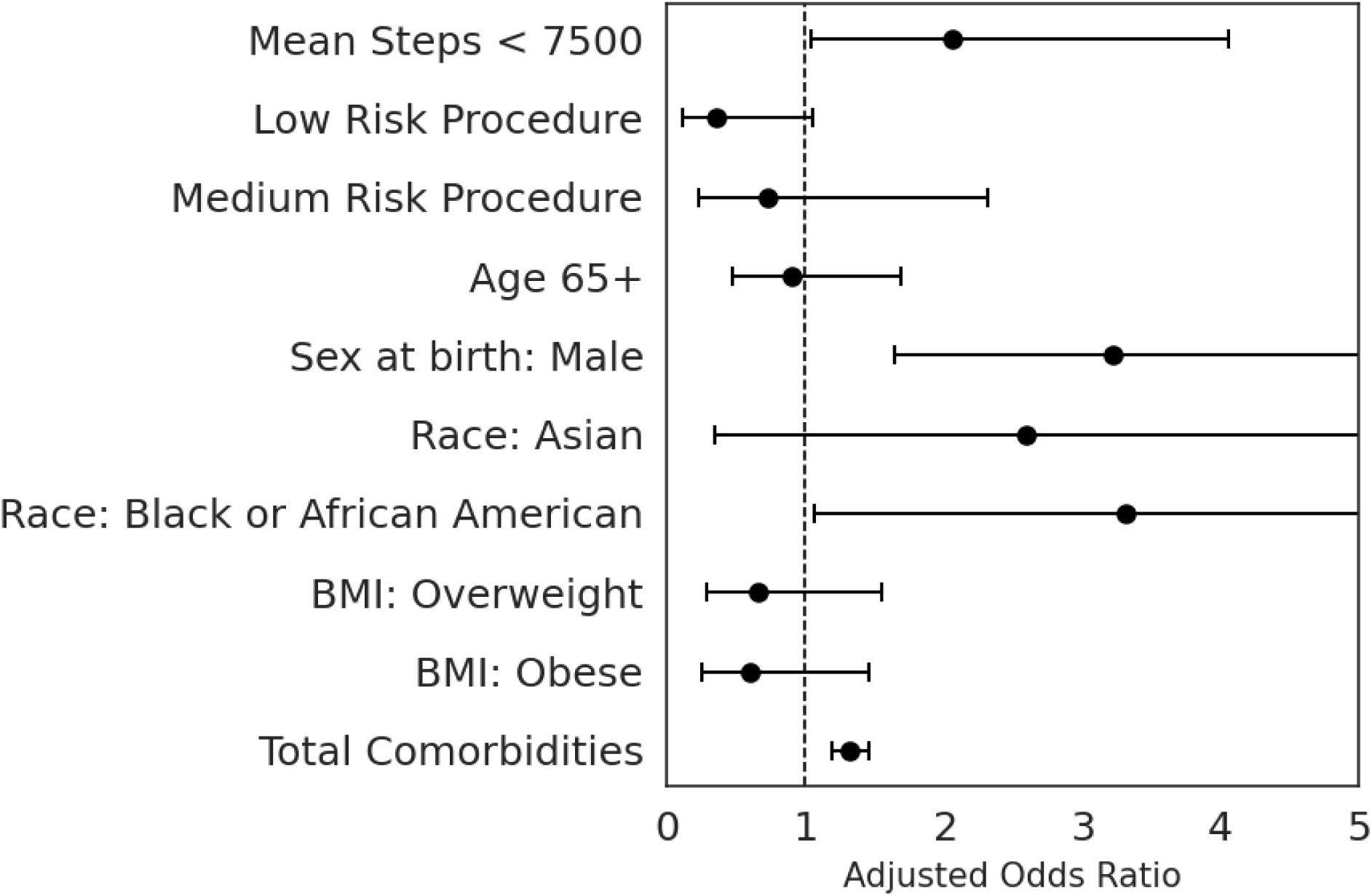
Association between average daily steps (<7,500) and development of adverse postoperative outcome based on multivariable logistic regression adjusting for clinical and demographic characteristics. Model diagnostics were performed with a C-index of 0.78, AIC of 320, and BIC of 362. BMI: body mass index.

**Figure 4:**
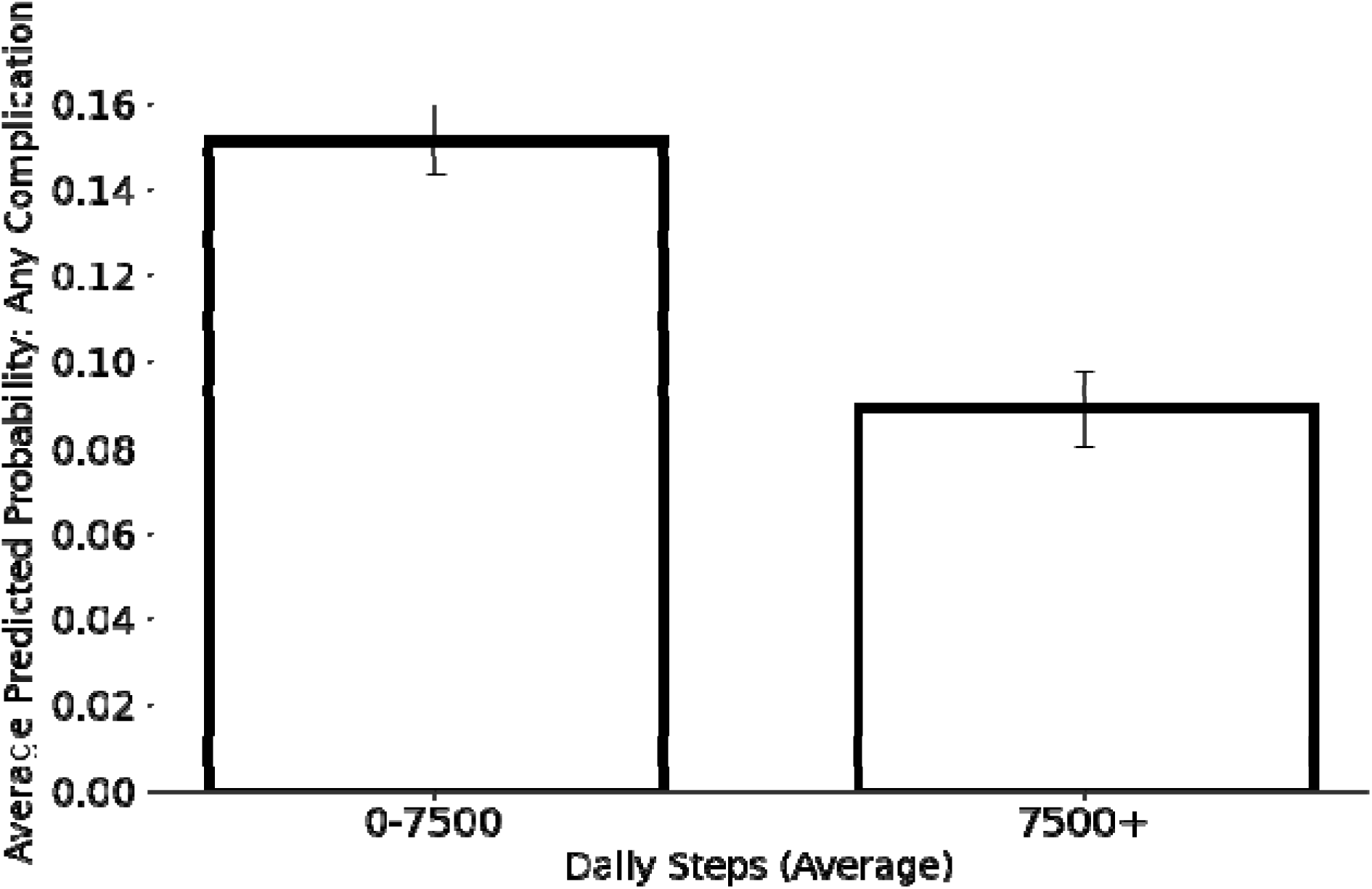
Predicted probability of any adverse event based on estimates from fitted logistic regression model (including procedural risk, age, sex, race, comorbid disease) in participants above and below 7,500-step threshold. Error bars denote standard error.

## DISCUSSION

In this retrospective observational cohort study of participants in the AOU Research Program, we found that preoperative daily step counts taken from EHR-linked wearable device data were an independent predictor for the development of adverse postoperative outcomes across a range of surgical procedures. After adjusting for patient demographic, clinical, and surgical factors, individuals who had an average daily step count of less than 7,500 steps prior to their procedure were at increased odds of developing adverse outcomes in the 90 days after surgery.

The results in this study are consistent with past studies that have examined postoperative outcomes in patients taking more or less than a set number of steps in the preoperative period.^13, 14^ The work reported here expands the scope of preoperative fitness assessment using wearable devices by utilizing a large and diverse surgical cohort, including several procedures of varying risk. The potential for using wearable devices to improve clinical care is supported by several prior studies in addition to ours. For example, after several surgical procedures, a higher step count on postoperative day one was associated with a decreased probability of a prolonged length of stay.^22^ In peritoneal cancer, a higher mean daily step count during the inpatient period predicted a lower risk of hospital readmission following resection.^23^ Others have shown that activity monitoring in the postoperative setting can help increase patient activity levels and ultimately improve outcomes.^24, 25^ In each of these studies, the use of wearable devices provided data to guide treatment that otherwise would not have been available through other monitoring tools.

Wearable devices can provide a more objective measure of preoperative fitness than patient-reported activity levels and a more cost-effective, efficient option than CPET. Wearable devices have previously been tested in the immediate preoperative period to provide similar information to CPET tests, such as 6-minute walking time.^26^ To translate wearable devices into clinical practice, a more rigorous selection of important features, their relative weights in determining surgical outcomes, and their interactions with other important clinical covariates should be further studied. For example, Cos et al. developed machine learning models that incorporated multiple domains of wearable device data to predict surgical outcomes in a cohort of pancreatectomy patients.^27^ Najafi et al. found a strong association between scores on a wrist-worn frailty-meter and major adverse events following lower extremity revascularization.^28^ Activity measured by wearable devices has also been shown to be associated with a shorter length of stay following major surgery.^22^

The growing body of work surrounding wearable devices has highlighted several areas in which they can improve care by replacing, adding to, and refining current measures of risk stratification. Additionally, several recent studies have shown patient fitness can be improved in the preoperative period through prehabilitation programs.^29, 30^ These programs can even be delivered through digital and virtual tools, such as smartwatches and mobile applications, making them accessible and easier to use for many patients.^31^ This suggests that wearable devices can not only be used for preoperative risk stratification, but also can be a mechanism to help improve postoperative outcomes.

Notable to this study was the use of the AOU Research Program to assess the association between preoperative step counts and surgical outcomes. The AOU Research Program presents a unique data source for conducting observational surgical research. Relative to other programs that have collected longitudinal health data, AOU has prioritized the recruitment of groups traditionally underrepresented in biomedical research to better reflect the diversity of the United States and make findings more generalizable to the population.^16^ Additionally, the AOU Research Program integrates data sources not typically available in other large datasets intended for biomedical research, such as wearable device, genetic, and participant-reported surveys.

In the context of the AOU Research Program and the integration of new technologies like wearable devices in healthcare, the importance of health equity cannot be overstated.^32^ Our findings underscore the necessity of making these technological advancements accessible and beneficial to the entire population, especially groups traditionally underrepresented in biomedical research. Future research should focus on understanding participants’ technological literacy, a crucial element for effectively using and interpreting wearable device data.^33^ Ensuring patients’ comfort and proficiency with these technologies is essential for enhancing their engagement and utilization. Additionally, preparing health providers to integrate this technology into their clinical practice, including the incorporation of wearable device data into EHR, is crucial. Exploring other implementation challenges and facilitators, such as addressing data privacy and managing cost considerations is also critical.^34^ Assessing the feasibility of implementing wearable technology in various healthcare settings will be key in identifying gaps and opportunities for ensuring equitable access and use of these technologies across diverse healthcare environments.^35^

The findings of this study must be considered in the context of the limitations. First, the step counts used as a preoperative measure did not reflect the immediate timeframe before surgery. Instead, all wearable device data that was generated by a patient prior to their surgery was factored into the average preoperative step count. As a result, the reported step count reflects chronic activity level and specific conclusions about how acute changes in step count leading up to surgery cannot be commented on based on the data available. Second, while AOU has focused on recruiting historically underrepresented groups in biomedical research, the included participants all had wearable device data from a personal Fitbit, and the study cohort may not be representative of a broader population. Third, there was heterogeneity among surgical procedures included in the study that included surgeries which may have treated conditions that could directly impact daily step counts (i.e., orthopedic procedures). However, the approach to include longer-term step count data allows an assessment of chronic activity that is less sensitive to the condition-specific changes that occurred closer to the time of surgery. For example, the average number of days of step data included was 851 (Q1: 240; Q3: 1343) per patient. Lastly, while AOU has sought to enroll a diverse group of participants, with many participants belonging to groups traditionally underrepresented in biomedical research, it relies on a bring-your-own-device (BYOD) model for collecting Fitbit data. BYOD programs have traditionally suffered from a lack of diversity, and this is evidenced by the study cohort being comprised of largely white, female, non-Hispanic or Latino participants.(31)

In conclusion, average daily steps are associated with elective surgical outcomes across a range of common procedures and the relationship is independent of several clinically relevant factors including comorbid conditions and type of surgery. The results of this study support the use of wearable device data and a step threshold of 7,500 steps as a guide to preoperative risk stratification and optimization programs prior to elective surgery.

## Data Availability

All data produced in the present work are contained in the manuscript.

## ACKNOWLEDGEMENTS

The All of Us Research Program is supported by the National Institutes of Health, Office of the Director: Regional Medical Centers: 1 OT2 OD026549; 1 OT2 OD026554; 1 OT2 OD026557; 1 OT2 OD026556; 1 OT2 OD026550; 1 OT2 OD 026552; 1 OT2 OD026553; 1 OT2 OD026548; 1 OT2 OD026551; 1 OT2 OD026555; IAA #: AOD 16037; Federally Qualified Health Centers: HHSN 263201600085U; Data and Research Center: 5 U2C OD023196; Biobank: 1 U24 OD023121; The Participant Center: U24 OD023176; Participant Technology Systems Center: 1 U24 OD023163; Communications and Engagement: 3 OT2 OD023205; 3 OT2 OD023206; and Community Partners: 1 OT2 OD025277; 3 OT2 OD025315; 1 OT2 OD025337; 1 OT2 OD025276. In addition, the All of Us Research Program would not be possible without the partnership of its participants.

## SUPPLEMENTARY DATA

**Supplementary Table 1:** CPT codes included in the NSQIP targeted procedures dataset were used as inclusion codes for creating a surgical cohort in AOU.

| Procedure Name | Surgery<br>N=475 (%) | CPT Codes |
| --- | --- | --- |
| Low Risk |  |  |
| **Abdominoplasty** | < 20 (< 4.2%) | 15830 |
| **Aortoiliac - Endo** | < 20 (< 4.2%) | 34707, 34708, 37220, 37221 |
| **Appendectomy** | < 20 (4.2%) | 44950, 44960, 44970 |
| **Bariatric** | 42 (8.8%) | 43644, 43645, 43770, 43773, 43775, 43842, 43843, 43845, 43846, 43847, 43848 |
| **Bladder Suspension** | < 20 (4.2%) | 57288 |
| **Breast Reconstruction** | 32 (6.7%) | 19325, 19340, 19342, 19357, 19361, 19364, 19367, 19368, 19369 |
| **Breast Reduction** | 22 (4.6%) | 19318 |
| **Carotid-Vertebral-Open** | < 20 (< 4.2%) | 34001, 35001, 35002, 35005, 35180, 35188, 35201, 35231, 35261, 35301, 35501, 35506, 35508, 35509, 35510, 35601, 35606, 35642, 35645, 35691, 35693, 35694, 35695 |
| **Gynecological Reconstruction** | < 20 (< 4.2%) | 57260, 57265, 57267, 57268, 57270, 57280, 57282, 57283 |
| **Hysterectomy/Myomectomy** | 72 (15.2%) | 58140, 58145, 58146, 58150, 58152, 58180, 58200, 58210, 58240, 58260, 58262, 58263, 58267, 58270, 58275, 58280, 58285, 58290, 58291, 58292, 58294, 58541, 58542, 58543, 58544, 58545, 58546, 58548, 58550, 58552, 58553, 58554, 58570, 58571, 58572, 58573, 58575, 58940, 58943, 58950, 58951, 58952, 58953, 58954, 58956 |
| **Lower Extremity (Infrainguinal)-Endo** | < 20 (< 4.2%) | 37224, 37225, 37226, 37227, 37228, 37229, 37230, 37231 |
| **Prostatectomy** | < 20 (< 4.2%) | 55801, 55810, 55812, 55815, 55821, 55831, 55840, 55842, 55845, 55866 |
| **Thyroidectomy** | 27 (5.7%) | 0200, 60210, 60212, 60220, 60225, 60240, 60252, 60254, 60260, 60270, 60271 |
| **Total Hip Arthroplasty** | 42 (8.8%) | 27125, 27130, 27132, 27134, 27137, 27138 |
| **Total Knee Arthroplasty** | 65 (13.7%) | 27447, 27486, 27487 |
| **Transurethral Resection of Prostate (TURP)** | < 20 (< 4.2%) | 52601, 52630, 52647, 52648, 52649 |
| Medium Risk |  |  |
| **Descending Thoracic Aorta-Endo** | < 20 (< 4.2%) | 33880, 33881, 33883, 33886, 33889, 33891 |
| **Flap** | < 20 (< 4.2%) | 15731, 15730, 15733, 15734, 15736, 15738, 15740, 15750, 15756, 15757, 15758 |
| **Lung Resection** | < 20 (< 4.2%) | 32440, 32442, 32445, 32480, 32482, 32484, 32486, 32488, 32491, 32503, 32504, 32505, |
|  |  | 32506, 32507, 32663, 32666, 32667, 32668, 32669, 32670, 32671, 32672 |
| **Nephrectomy** | < 20 (< 4.2%) | 50220, 50225, 50230, 50234, 50236, 50240, 50543, 50545, 50546, 50548 |
| **Spine** | 61 (12.8%) | 22100, 22101, 22102, 22110, 22112, 22114, 22206, 22207, 22210, 22212, 22214, 22220, 22222, 22224, 22318, 22319, 22325, 22326, 22327, 22526, 22532, 22533, 22548, 22551, 22554, 22556, 22558, 22586, 22590, 22595, 22600, 22610, 22612, 22630, 22633, 22800, 22802, 22804, 22808, 22810, 22812, 22818, 22819, 22830, 22849, 22850, 22852, 22855, 22856, 22857, 22858, 22860, 22861, 22862, 22864, 22865, 22867, 22869, 63001, 63003, 63005, 63011, 63012, 63015, 63016, 63017, 63020, 63030, 63040, 63042, 63045, 63046, 63047, 63050, 63051, 63055, 63056, 63064, 63075, 63077, 63081, 63085, 63087, 63090, 63101, 63102, 63170, 63172, 63173, 63185, 63190, 63191, 63197, 63200, 63250, 63251, 63252, 63265, 63266, 63267, 63268, 63270, 63271, 63272, 63273, 63275, 63276, 63277, 63278, 63280, 63281, 63282, 63283, 63285, 63286, 63287, 63290, 63300, 63301, 63302, 63303, 63304, 63305, 63306, 63307 |
| **Upper Extremity-Open** | < 20 (< 4.2%) | 34101, 34111, 35011, 35013, 35045, 35206, 35236, 35266, 35321, 35511, 35512, 35516, 35518, 35522, 35523, 35525, 35612, 35616, 35650 |
| High Risk |  |  |
| **Abdominal (Infrarenal) Aorta-Open** | < 20 (< 4.2%) | 34830, 34831, 34832, 35081, 35082, 35091, 35092, 35102, 35103 |
| **Aortic Arch & Proximal Brachiocephalic Vessels-Open** | < 20 (< 4.2%) | 34051, 35021, 35022, 35311, 35526, 35626 |
| **Brain Tumor** | < 20 (< 4.2%) | 61510, 61512, 61518, 61519, 61520, 61521, 61526, 61530, 61545, 61546 |
| **Colectomy** | < 20 (< 4.2%) | 44140, 44141, 44143, 44144, 44145, 44146, 44147, 44150, 44151, 44160, 44204, 44205, 44206, 44207, 44208, 44210 |
| **Cystectomy** | < 20 (< 2.4%) | 51550, 51555, 51565, 51570, 51575, 51580, 51585, 51590, 51595, 51596, 51597 |
| **Esophagectomy** | < 20 (< 2.4%) | 43101, 43107, 43108, 43112, 43113, 43116, 43117, 43118, 43121, 43122, 43123, 43124, 43286, 43287, 43288 |
| **Hepatectomy** | < 20 (< 4.2%) | 47120, 47122, 47125, 47130 |
| **Hip Fracture** | < 20 (< 4.2%) | 27236, 27244, 27245 |
| **Lower Extremity (Infrainguinal)-Open** | < 20 (< 4.2%) | 34201, 34203, 35141, 35142, 35151, 35152, 35226, 35286, 35302, 35303, 35304, 35305, |
|  |  | 35371, 35372, 35556, 35566, 35570, 35571, 35583, 35585, 35587, 35656, 35666, 35671, 35879, 35881, 35883, 35884 |
| **Pancreatectomy** | < 20 (< 4.2%) | 48120, 48140, 48145, 48146, 48148, 48150, 48152, 48153, 48154, 48155, 48999 |
| **Proctectomy** | < 20 (< 4.2%) | 44155, 44156, 44157, 44158, 44211, 44212, 45110, 45111, 45112, 45113, 45114, 45116, 45119, 45120, 45121, 45123, 45126, 45130, 45135, 45160, 45395, 45397, 45402, 45550 |
| **Visceral Vessels-Open** | < 20 (< 4.2%) | 34151, 35111, 35112, 35121, 35122, 35341, 35531, 35535, 35536, 35560, 35631, 35632, 35633, 35634, 35636 |

**Supplementary Table 2:** Adverse postoperative events were defined using SNOMED codes.

| **Postoperative Complications** | **SNOMED Codes** |
| --- | --- |
| Acute Respiratory Distress Syndrome | 67782005 |
| Cardiac Arrhythmia | 698247007 |
| Deep Venous Thrombosis | 128053003 |
| Pulmonary Embolism | 59282003 |
| Pneumonia | 233604007 |
| Renal Failure | 42399005 |
| Respiratory Failure | 409622000 |
| Sepsis | 91302008 |
| Surgical Site Infection | 609340004, 762611002, 609339001, 433202001 |
| Urinary Tract Infection | 68566005 |

**Supplementary Table 3:** Baseline characteristics of the study cohort separated by active vs. Inactive group.

|  | **> 7500 steps/day**<br>**(N = 191)** | **< 7500 steps/day**<br>**(N = 284)** | **P** |
| --- | --- | --- | --- |
| Age, years |  |  |  |
| **Median, (Q1-Q3)** | 59.7 (48.4, 66.4) | 58.8 (47.8, 68.2) | 0.87 |
| **18-29** | < 20 (< 10.5%) | < 20 (< 7.0%) |  |
| **30-49** | 50 (26.2%) | 71 (25.0%) | 0.77 |
| **50-64** | 70 (36.6%) | 104 (36.6%) | 0.99 |
| **65+** | > 51 (> 26.7%) | > 89 (> 31.3%) |  |
| Sex at birth |  |  |  |
| **Female** | > 118 (> 61.8%) | > 209 (> 73.6%) |  |
| **Male** | 53 (27.7%) | 55 (19.4%) | 0.03 |
| **Missing or Unknown** | < 20 (< 10.5%) | < 20 (< 7.0%) |  |
| Race |  |  |  |
| **White** | 163 (85.3%) | 242 (85.2%) | 0.97 |
| **Non-White or Unknown** | 28 (14.7%) | 42 (14.8%) | 0.97 |
| Ethnicity |  |  |  |
| **Not Hispanic or Latino** | > 177 (> 92.7%) | 256 (90.1%) |  |
| **Hispanic or Latino or Unknown** | < 20 (<10.5%) | 28 (9.9%) |  |
| Elixhauser Comorbidity Index |  |  |  |
| **0** | 36 (18.8%) | 31 (10.9%) | 0.01 |
| **1** | 31 (16.2%) | 25 (8.8%) | 0.01 |
| **2** | 31 (16.2%) | 49 (17.3%) | 0.77 |
| **3** | 26 (13.6%) | 38 (13.4%) | 0.94 |
| **4+** | 67 (35.1%) | 141 (49.6%) | 0.002 |
| Body Mass Index (BMI) |  |  |  |
| **Median (Q1, Q3)** | 27.3 (24.3, 31.6) | 31.3 (26.8, 38.5) | < 0.001 |
| Comorbidities |  |  |  |
| **Hypertension, Uncomplicated** | 75 (39.3%) | 135 (47.5%) | 0.075 |
| **Obesity** | 43 (22.5%) | 123 (43.3%) | < 0.001 |
| **Solid Tumor Without Metastasis** | 64 (33.5%) | 78 (27.5%) | 0.16 |
| **Depression** | 37 (19.3%) | 102 (35.9%) | < 0.001 |
| **Chronic Pulmonary Disease** | 40 (20.9%) | 85 (29.9%) | 0.029 |
| **Hypothyroidism** | 26 (13.6%) | 70 (24.6%) | 0.0033 |
| **Diabetes,**<br>**Uncomplicated** | 22 (11.5%) | 45 (15.8%) | 0.18 |
| **Liver Disease** | 24 (12.6%) | 35 (12.3%) | 0.94 |
| Relative Risk of<br>Surgery |  |  |  |
| **Low Risk** | 143 (74.9%) | 216 (76.1%) | 0.77 |
| **Medium Risk** | > 28 | > 48 |  |
| **High Risk** | < 20 | < 20 |  |

**Supplementary Figure 1:**
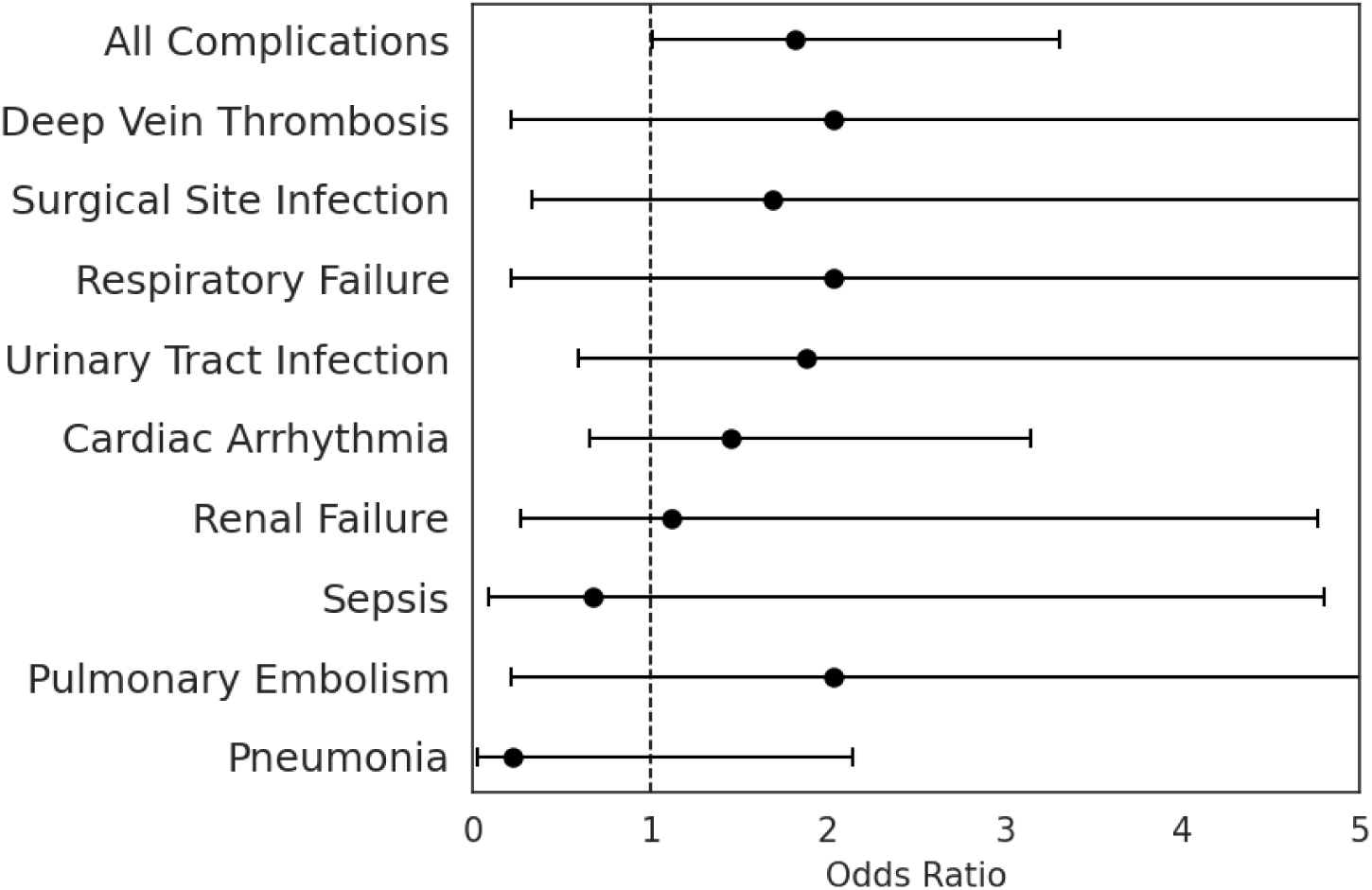
Unadjusted analysis was performed on all postoperative adverse events and each individual event between active and inactive groups (odds ratios showing inactive relative to active). Any codes indicating chronic conditions were excluded.

## REFERENCES

1 Moonesinghe SR, Mythen MG, Das P et al. Risk Stratification Tools for Predicting Morbidity and Mortality in Adult Patients Undergoing Major Surgery: Qualitative Systematic Review. Anesthesiology 2013; 119 (4): 959–981.

2 Cole WW, Familia M, Miskimin C, Mulcahey MK. Preoperative Optimization and Tips to Avoiding Surgical Complications Before the Incision. Sports Medicine and Arthroscopy Rev 2022; 30 (1): 2–9.

3 Bhagwat M, Paramesh K. Cardio-pulmonary exercise testing: An objective approach to pre-operative assessment to define level of perioperative care. Indian J Anaesth 2010; 54 (4): 286–291.

4 Silvapulle E, Darvall J. Subjective methods for preoperative assessment of functional capacity. BJA Education 2022; 22 (7): 249–257.

5 Molenaar CJL, Papen-Botterhuis NE, Herrle F, Slooter GD. Prehabilitation, making patients fit for surgery - a new frontier in perioperative care. Innovative Surgical Sciences 2019; 4 (4): 132–138.

6 Chen BP, Awasthi R, Sweet SN et al. Four-week prehabilitation program is sufficient to modify exercise behaviors and improve preoperative functional walking capacity in patients with colorectal cancer. Supportive Care in Cancer 2017; 25 (1): 33–40.

7 Minnella EM, Awasthi R, Loiselle S-E et al. Effect of Exercise and Nutrition Prehabilitation on Functional Capacity in Esophagogastric Cancer Surgery: A Randomized Clinical Trial. JAMA Surg 2018; 153 (12): 1081–1089.

8 Fernández-Costa D, Gómez-Salgado J, Castillejo del Río A et al. Effects of Prehabilitation on Functional Capacity in Aged Patients Undergoing Cardiothoracic Surgeries: A Systematic Review. Healthc 2021; 9 (11): 1602.

9 Older PO, Levett DZH. Cardiopulmonary Exercise Testing and Surgery. Annals of the American Thorac Society 2017; 14 (Supplement_1): S74–S83.

10 Bailey DM, Halligan CL, Davies RG et al. Subjective assessment underestimates surgical risk: On the potential benefits of cardiopulmonary exercise testing for open thoracoabdominal repair. J Card Surg 2022; 37 (8): 2258–2265.

11 Wijeysundera DN, Pearse RM, Shulman MA et al. Assessment of functional capacity before major non-cardiac surgery: an international, prospective cohort study. The Lancet 2018; 391 (10140): 2631–2640.

12 Billé A, Buxton J, Viviano A et al. Preoperative Physical Activity Predicts Surgical Outcomes Following Lung Cancer Resection. Integr Cancer Therapies 2021; 20: 1534735420975853.

13 Hedrick TL, Hassinger TE, Myers E et al. Wearable Technology in the Perioperative Period: Predicting Risk of Postoperative Complications in Patients Undergoing Elective Colorectal Surgery. Diseases of the Colon & Rectum 2020; 63 (4): 538–544.

14 Nakajima H, Yokoyama Y, Inoue T et al. How Many Steps Per Day are Necessary to Prevent Postoperative Complications Following Hepato-Pancreato-Biliary Surgeries for Malignancy? Annals of Surgical Oncology 2020; 27 (5): 1387–1397.

15 Richards SJG, Jerram PM, Brett C et al. The association between low pre-operative step count and adverse post-operative outcomes in older patients undergoing colorectal cancer surgery. Perioperative Medicine 2020; 9 (1): 20.

16 Investigators AoURP. The “All of Us” Research Program. New Engl. J. of Medicine. 381. 2019:668–676.

17 Evenson KR, Goto MM, Furberg RD. Systematic review of the validity and reliability of consumer-wearable activity trackers. Int J Behav Nutr Phys Act 2015; 12: 159.

18 Bai Y, Tompkins C, Gell N et al. Comprehensive comparison of Apple Watch and Fitbit monitors in a free-living setting. PLoS One 2021; 16 (5): e0251975.

19 Verhagen NB, Koerber NK, Szabo A et al. Vaccination Against SARS-CoV-2 Decreases Risk of Adverse Events in Patients who Develop COVID-19 Following Cancer Surgery. Annals of Surgical Oncology 2023; 30 (3): 1305–1308.

20 Lee IM, Shiroma EJ, Kamada M et al. Association of Step Volume and Intensity With All-Cause Mortality in Older Women. JAMA Intern Med 2019; 179 (8): 1105–1112.

21 Tudor-Locke C, Craig CL, Brown WJ et al. How many steps/day are enough? For adults. Int J Behav Nutr Phys Act 2011; 8: 79.

22 Daskivich TJ, Houman J, Lopez M et al. Association of Wearable Activity Monitors With Assessment of Daily Ambulation and Length of Stay Among Patients Undergoing Major Surgery. JAMA Netw Open 2019; 2 (2): e187673–e187673.

23 Low CA, Bovbjerg DH, Ahrendt S et al. Fitbit step counts during inpatient recovery from cancer surgery as a predictor of readmission. Annals of Behavioral Medicine 2017; 52 (1): 88–92.

24 Ni C-y, Wang Z-h, Huang Z-p, et al. Early enforced mobilization after liver resection: A prospective randomized controlled trial. International J of Surgery 2018; 54: 254–258.

25 Wolk S, Linke S, Bogner A et al. Use of Activity Tracking in Major Visceral Surgery—the Enhanced Perioperative Mobilization Trial: a Randomized Controlled Trial. Journal of Gastroint Surg 2019; 23 (6): 1218–1226.

26 Greco M, Angelucci A, Avidano G et al. Wearable Health Technology for Preoperative Risk Assessment in Elderly Patients: The WELCOME Study. Diagnostics 2023; 13 (4): 630.

27 Cos H, Li D, Williams G et al. Predicting Outcomes in Patients Undergoing Pancreatectomy Using Wearable Technology and Machine Learning: Prospective Cohort Study. J Medical Internet Res 2021; 23 (3): e23595.

28 Najafi B, Veranyan N, Zulbaran-Rojas A et al. Association Between Wearable Device– Based Measures of Physical Frailty and Major Adverse Events Following Lower Extremity Revascularization. JAMA Netw Open 2020; 3 (11): e2020161–e2020161.

29 Steffens D, Young J, Beckenkamp PR et al. Feasibility and acceptability of a preoperative exercise program for patients undergoing major cancer surgery: results from a pilot randomized controlled trial. Pilot and Feasibility Stud 2021; 7 (1): 27.

30 Hall DE, Youk A, Allsup K et al. Preoperative Rehabilitation Is Feasible in the Weeks Prior to Surgery and Significantly Improves Functional Performance. J of Frailty & Aging 2023; 12 (4): 267–276.

31 Waller E, Sutton P, Rahman S et al. Prehabilitation with wearables versus standard of care before major abdominal cancer surgery: a randomised controlled pilot study (trial registration: NCT04047524). Surgical Endoscopy 2022; 36 (2): 1008–1017.

32 Canali S, Schiaffonati V, Aliverti A. Challenges and recommendations for wearable devices in digital health: Data quality, interoperability, health equity, fairness. PLOS Digital Health 2022; 1 (10): e0000104.

33 Richardson S, Lawrence K, Schoenthaler AM, Mann D. A framework for digital health equity. npj Digital Medicine 2022; 5 (1): 119.

34 Holko M, Litwin TR, Munoz F et al. Wearable fitness tracker use in federally qualified health center patients: strategies to improve the health of all of us using digital health devices. npj Digital Medicine 2022; 5 (1): 53.

35 Huhn S, Axt M, Gunga H-C et al. The Impact of Wearable Technologies in Health Research: Scoping Review. JMIR Mhealth Uhealth 2022; 10 (1): e34384.

